# A room of one’s own: separate accommodation after perinatal loss and the value of midwifery care

**DOI:** 10.64898/2026.07.29.26359245

**Authors:** Claudia Ravaldi, Alfredo Vannacci

## Abstract

**Problem:** Guidelines ask that a woman whose baby has died be cared for away from the sights and sounds of newborns, but they do not say where she should be cared for instead.

**Background:** One common way to move a bereaved woman away from newborns is to admit her to a gynaecology ward, although this could undermine midwifery care. To date, no study assessed the impact of bereavement space on grief or mental health.

**Aim:** To test whether separate accommodation is associated with better outcomes and reported care, and whether achieving separation in a gynaecology ward rather than an obstetric ward is associated with a difference in midwifery care.

**Methods:** Cross-sectional analysis of the Italian OPALE observatory. Of 2601 women reporting a perinatal loss, 1662 reported both their ward and whether they shared accommodation, giving four care configurations. Measures were the Perinatal Grief Scale, the NSESSS, satisfaction with accommodation, respectful care, ratings of the midwife and the nurse, and an indicator of no midwifery care, adjusted for type of loss, gestational age, maternal age, time since loss and geographical area.

**Findings:** Overall, 35.7% of women shared accommodation with mothers and newborns, falling from 47.1% of losses before 2015 to 26.9% from 2023 onwards. Sharing was associated with higher grief (adjusted difference 3.22, 95% CI 0.81 to 5.63), more post-traumatic stress symptoms (1.13, 0.17 to 2.09), lower satisfaction with accommodation (−30.05, −33.08 to −27.02) and less respectful care (−0.51 on 0 to 4, −0.64 to −0.37). Among women for whom separation had been achieved, those in a gynaecology ward were more satisfied with their accommodation (7.33, 3.85 to 10.82) but reported lower midwifery presence (−7.13, −11.18 to −3.09) and three times the odds of receiving no midwifery care (OR 3.05, 1.49 to 6.23), with no difference in respectful care or satisfaction.

**Discussion:** Relocation to gynaecology delivers the room the guidelines ask for but erodes the midwifery care they assume will accompany it, a deficit invisible in the woman’s global satisfaction.

**Conclusion:** Guidance requiring separate accommodation should add that separation must not remove the woman from midwifery care. A separate room within the obstetric ward satisfies both, and provided the better care in our sample.

**Statement of significance:** *Problem or issue:* Every major guideline on bereavement care asks that a woman whose baby has died not be cared for among newborns, yet the recommendation rests on consensus rather than measured outcomes, and it says nothing about where she should be cared for instead.

*What is already known:* Adherence to bereavement guidelines as a whole is associated with lower grief and post-traumatic stress symptoms. The specific contribution of physical space has not been quantified, and the professional consequences of moving a bereaved woman out of the maternity area are unknown.

*What this paper adds:* Separate accommodation is associated with lower grief, fewer post-traumatic stress symptoms and better reported care. Obtaining that separation by relocation to a gynaecology ward is associated with a measurable loss of midwifery presence and a threefold increase in the odds of receiving no midwifery care at all, which the woman’s overall satisfaction does not reveal.

## 1. Introduction

Around 2.6 million pregnancies a year end in stillbirth, and many more in miscarriage, termination for fetal anomaly or neonatal death. Bereaved parents carry a well-documented burden of grief, depression and post-traumatic stress.^1^ The main international guidelines on bereavement care converge on a small set of recommendations: honest communication, respect for the baby, informed choices about birth, the opportunity to create memories, structured aftercare, and care delivered in a quiet and private space, away from the sights and sounds of other women giving birth and of newborn babies.^2,3^

Evidence that adherence to these recommendations matters clinically is recent. In the CLASS study, women whose care satisfied more than 40 of 60 checklist items reported greater satisfaction, more respectful care and substantially lower grief and post-traumatic stress symptoms.^4^ Adherence was scored as a total across the checklist, so which recommendations were met, and by what means, is yet to be explored.

The recommendation on proper space and accommodation is universally endorsed in guidelines, but almost entirely unstudied. A 2025 narrative review of dedicated bereavement rooms found that fewer than a quarter of eligible articles even mentioned them, and that no study evaluated implementation, design standards or measurable impact on grief, mental health or family outcomes.^5^ Systematic reviews of interventions to reduce distress after stillbirth reach the same conclusion: the few controlled studies that exist are at high risk of bias and the certainty of evidence is very low for every outcome.^6^ The qualitative record is consistent: parents describe the cheerful, bustling atmosphere of the labour ward as a painful place to be after their baby has died,^7^ name adequate time and appropriate physical space among the central themes of their account of hospital care,^8^ and ask for privacy without abandonment.^9^ The recommendation therefore rests on what parents report, and has never been tested against a measured outcome. It is also left unspecified, so where a woman is accommodated often depends on what is free on the day rather than on a rule.

The wider literature on hospital accommodation does not fill the gap. Outside intensive care, a systematic review of 145 papers on single-occupancy accommodation could draw no consistent conclusion about clinical benefit, and a minority of patients prefer shared accommodation because company protects them from loneliness.^10^ In maternity settings, single-room care is associated with higher maternal satisfaction on a weak quantitative evidence base.^11,12^ In that literature the expected benefits are quieter surroundings, more control and more dignity. After a perinatal death the exposure is the presence of other newborns, and no study has measured whether removing this exposure changes the women’s reports or their wellbeing.

Guidelines are quite clear on where the woman should not be, but they say little about where she should be, and nothing about who should care for her once she is moved. In a maternity service organised around birth, the simplest way to move a bereaved woman away from newborns is to admit her to the gynaecology ward, and this is often done with evident good intent. Midwives, however, are generally rostered to obstetric areas and deployed according to birth activity, while gynaecology wards are staffed predominantly by nurses whose routine work is not maternity care. Moving her may therefore remove the professional group whose scope of practice covers the care she still needs, since a woman who has given birth requires midwifery care whatever the outcome of her pregnancy. Midwifery staffing and skill mix are not interchangeable with total staffing.^13,14^ Furthermore, bereavement care is a specific competence: in a national survey of Italian midwives almost all underlined the importance of specific training, focusing on the hardness of communicating the loss, and of the clinical management that follows a death, including inhibition of lactation.^15,16^

Italy had no national guidance on the management of intrauterine death until the scientific societies issued recommendations in 2023.^17^ Care varied widely between units,^18^ and women were not always involved in the decisions that followed a stillbirth.^19^ The CLASS validation analysis used this observatory’s earlier recruitment, from the years before those recommendations appeared: it documented the consequences of their absence and argued that national guidance was needed.^4^ The present work extends the same observatory with losses now spannig the years before and after the recommendations.

The aims of this study were: to estimate the proportion of women in Italy who share their accommodation with mothers and newborn babies after a perinatal loss; to explore whether separate accommodation is associated with lower grief, fewer post-traumatic stress symptoms and better reported care; and to assess whether achieving separation in a gynaecology ward rather than in an obstetric ward is associated with a difference in the midwifery care women report. We also describe change over time in ward placement and in the space-related checklist items, taking the observatory’s earlier recruitment, the years covered by the CLASS validation analysis, as the pre-recommendation reference.

## 2. Methods

### 2.1 Design and setting

This is a cross-sectional analysis of OPALE (Observatory on PerinatAL hEalth), a continuously recruiting national online survey hosted on Qualtrics and run by the PeaRL laboratory of the University of Florence with the CiaoLapo Foundation. The survey is distributed through the Foundation’s channels and is open to women with any experience of pregnancy, with or without loss, and to any time since the event. Information was provided on the first page and consent obtained before the questionnaire began.

### 2.2 Participants and samples

We included women who reported a perinatal loss, defined as stillbirth, neonatal death, miscarriage or termination of pregnancy for fetal or maternal pathology, and who were admitted to hospital. Of 2601 such women, 1769 answered the item on shared accommodation and were used for the analysis of prevalence, of change over time and of the association between accommodation and outcomes. Of these, a further 107 were excluded because the ward was other than obstetric or gynaecology, was a combined unit, or was unknown, leaving 1662 women with a defined care configuration. A pre-specified sensitivity analysis was restricted to the 963 women whose loss occurred at 20 weeks or later, which reproduces the inclusion criterion used in the earlier CLASS analysis^4^ and keeps the two periods comparable.

The comparison over time is internal to the observatory. Its earlier recruitment, which supplied the CLASS analysis, contributed 261 women, all with a stillbirth at 20 weeks or later and with losses occurring between 2008 and 2019, entirely before the 2023 recommendations. The women recruited since contribute the corresponding stratum of 736 women with a stillbirth at 20 weeks or later, of whom 304 had a loss from 2023 onwards. The checklist and the recruitment channels are the same throughout, and the two groups are distinguished only by when the women answered.

### 2.3 Measures

#### Exposure

Women were asked whether they spent their admission in a room with mothers and their newborn babies (yes, no, do not remember), and what type of ward they were admitted to. Crossing the two produced four configurations: obstetric or gynaecology ward, each with separate or shared accommodation. Ward was recovered from free-text responses where the closed question had been answered “other” but the text named a ward unambiguously; the closed question always took precedence. The item establishes that the woman was not accommodated with mothers and newborns, not necessarily she was in a single room.

#### Midwifery presence

Two matched item batteries asked how important each of the obstetrician, the midwife, the nurse and the psychologist had been for the woman’s medical and clinical care and, separately, for her emotional and relational care, rated from 0 to 100, with a “not applicable” option for professionals she was not cared for by. From these we derived the rating given to the midwife; a within-woman skill-mix differential, the midwife rating minus the nurse rating, which removes a woman’s general tendency to rate care highly or poorly and separates the composition of care from its overall quality; and an indicator of no midwifery care, defined as having rated other figures, but not the midwife.

#### Experience, reported care and psychological outcomes

Satisfaction with hospital accommodation was rated from 0 to 100. Two items adapted from the instrument developed by Bohren and colleagues to measure how women are treated during childbirth^20^ asked whether the woman had been treated with respect and whether she was satisfied overall with the services received, each scored 0 to 4. Grief was measured with the 33-item short version of the Perinatal Grief Scale,^21^ in the Italian translation validated within this observatory,^22^ and post-traumatic stress symptoms with the National Stressful Events Survey PTSD Short Scale.^23^ For the subsample completing the CLASS checklist^4^ we used the ward-environment section and three individual items: the offer of the least disruptive ward and, if possible, a single room; the assurance of parents’ privacy; and the choice of a quiet environment for difficult conversations. The whole 60-item score was retained as a contrast, to establish whether any change over time was specific to space or general.

#### Time

Year of loss was grouped into four periods (before 2015, 2015 to 2019, 2020 to 2022, 2023 onwards) and also used as a binary indicator of loss from 2023 onwards, the year the recommendations were issued.

### 2.4 Analysis

We report means with 95% confidence intervals and proportions by configuration. Differences were estimated by linear regression, or logistic regression for binary indicators, adjusted for type of loss, gestational age at loss, maternal age at loss, years elapsed since the loss and geographical area of residence. For accommodation and outcomes the comparison was between separate and shared accommodation, with a further model adding ward type and a test of the interaction between the two. For midwifery presence the obstetric ward with separate accommodation was the reference, and the comparison of primary interest was with the gynaecology ward with separate accommodation, since that contrast isolates the consequence of how separation was obtained rather than whether it was obtained. For change over time we fitted the four-period model, then a model containing both the indicator for 2023 onwards and year of loss as a continuous term, so that the step could be assessed against a smooth secular trend. Analyses were complete-case, in Stata 18 and Python 3.

## 3. Results

### 3.1 Sample and prevalence of shared accommodation

The 1662 women included had experienced a stillbirth, a miscarriage, a termination of pregnancy for fetal anomaly or a neonatal death, and had been admitted either to an obstetric or to a gynaecology ward. Groups were well balanced on education (p=0.995) and on previous perinatal loss (p=0.082), and differed on gestational age, type of loss, time since the loss and geographical area, all of which were adjusted for. The four configurations were unevenly distributed: 546 women (32.9%) were in an obstetric ward with separate accommodation, 405 (24.4%) in an obstetric ward with shared accommodation, 519 (31.2%) in a gynaecology ward with separate accommodation and 192 (11.6%) in a gynaecology ward with shared accommodation. Separation was achieved for 57.4% of women admitted to an obstetric ward and for 73.0% of those admitted to gynaecology. A full description of the sample is given in Table S1.

Among all women who answered the accommodation item, 632 of 1769 (35.7%) spent their admission alongside mothers and newborns. Prevalence varied markedly by type of loss: 53.6% after neonatal death, 44.6% after miscarriage, 31.5% after termination for fetal anomaly and 25.2% after stillbirth (p<0.001). The lowest figure was for the one event for which national recommendations exist, the highest for events they do not cover. Prevalence also varied geographically, from 29.5% in the North to 52.7% in the South (p<0.001).

The CLASS item on ward placement showed high adherence in both separated groups (84.0 and 84.5) and low adherence in both shared groups (58.3 and 47.9; p<0.001).

### 3.2 A room away from newborns: accommodation and outcomes

Women who shared their accommodation with mothers and newborns reported higher grief (Perinatal Grief Scale 103.5 versus 101.8; adjusted difference 3.22, 95% CI 0.81 to 5.63, p=0.009) and more post-traumatic stress symptoms (15.9 versus 15.0; adjusted difference 1.13, 95% CI 0.17 to 2.09, p=0.021). Satisfaction with accommodation was 38.0 among women who shared and 72.5 among those who did not (adjusted difference −30.05, 95% CI −33.08 to −27.02, p<0.001). On the two items scored 0 to 4, women who shared were less likely to report having been treated with respect (2.72 versus 3.36; −0.51, 95% CI −0.64 to −0.37) and less satisfied overall with the services they received (2.12 versus 3.01; −0.72, 95% CI −0.88 to −0.55, both p<0.001). In the checklist subsample, adherence on the ward-environment section was 52.4 among women who shared and 78.3 among those who did not (−20.07, 95% CI −26.58 to −13.56, p<0.001), with differences of similar magnitude on the individual items on privacy and on a quiet environment. Table 1 shows the means by accommodation, Figure 1 the adjusted differences and Table S2 their confidence intervals.

**Table 1.**
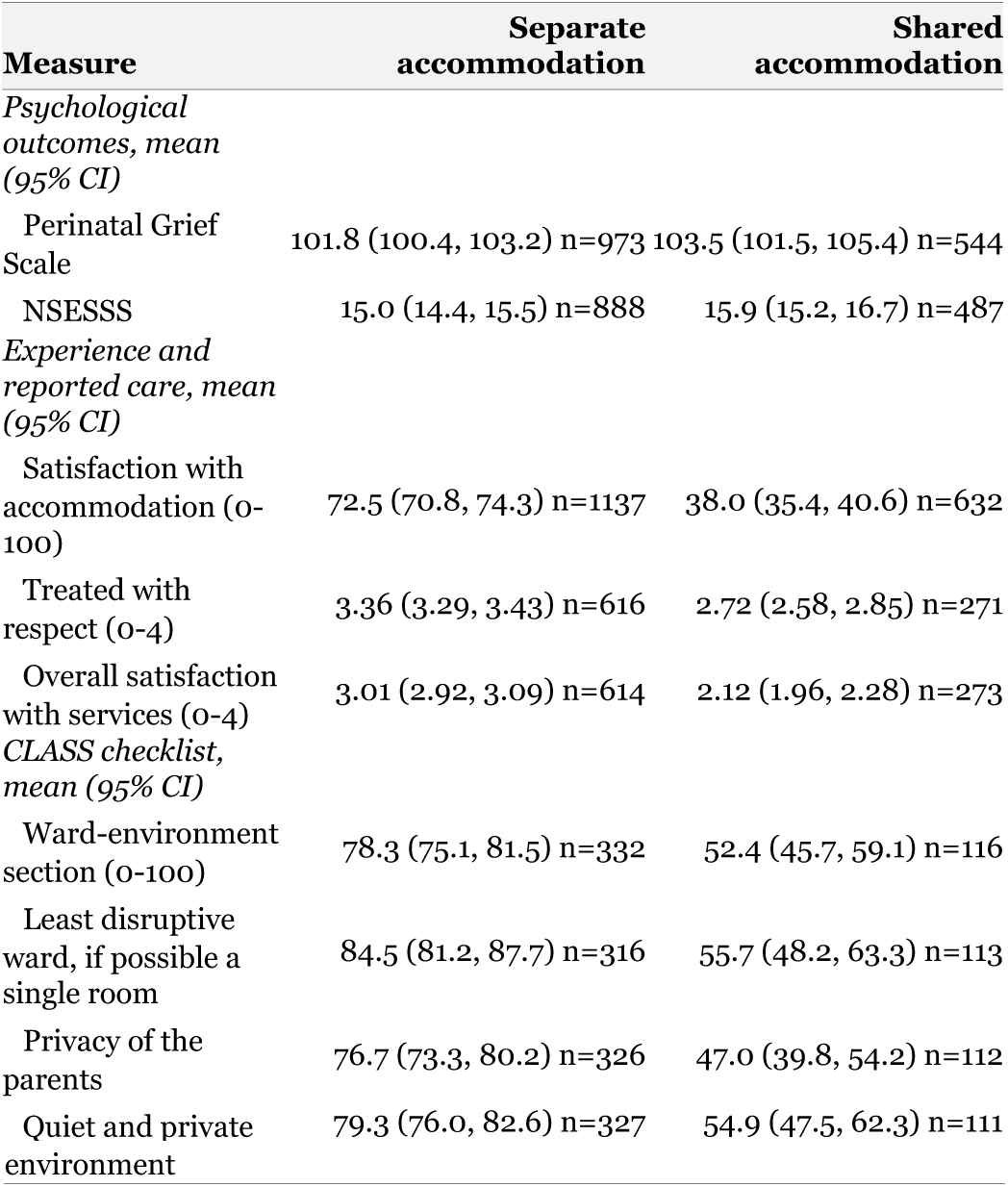
Psychological outcomes, reported care and checklist adherence, by whether accommodation was shared with mothers and newborn babies.

**Figure 1.**
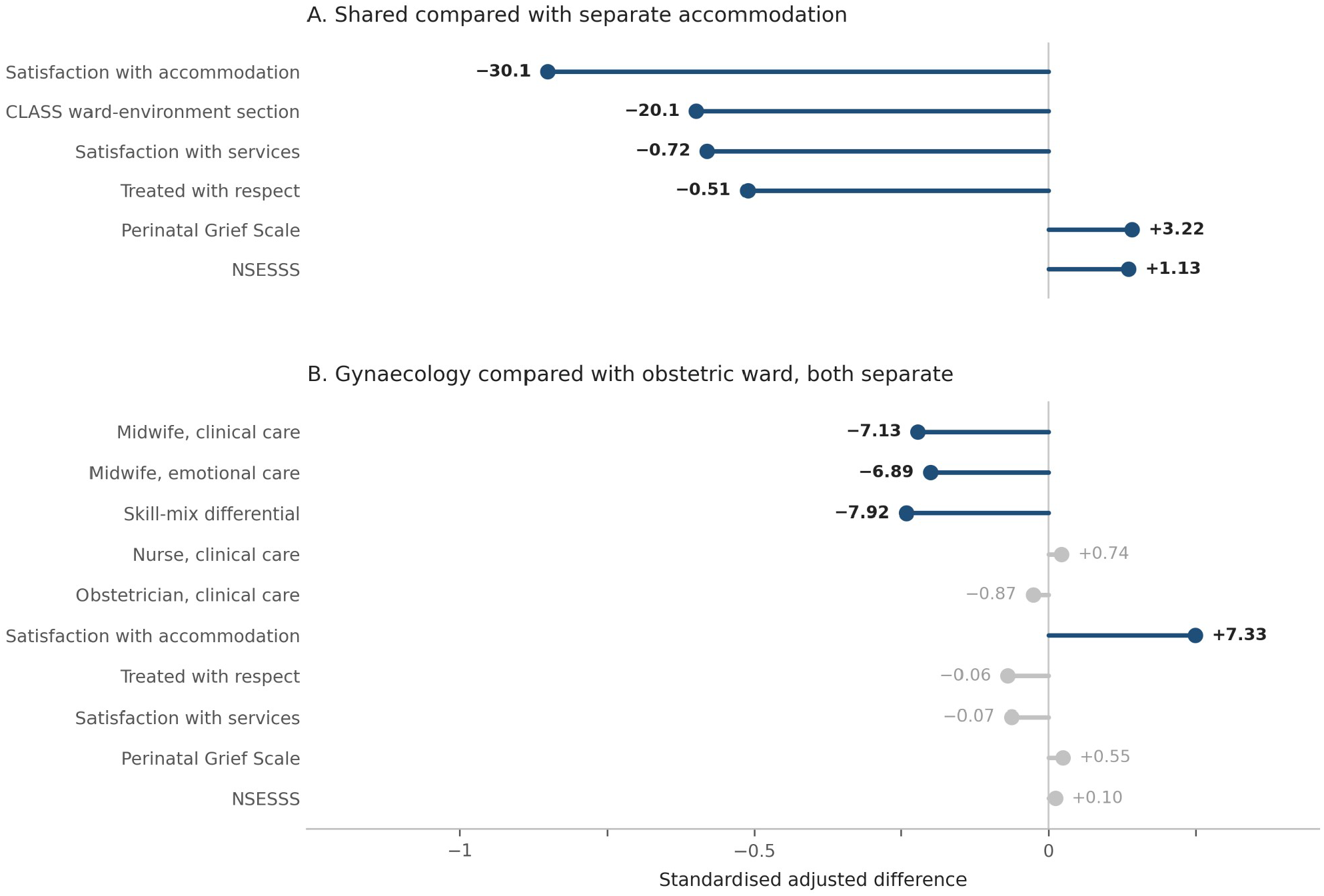
Adjusted differences. Panel A compares shared with separate accommodation, panel B the gynaecology with the obstetric ward among women with separate accommodation. Position on the axis is the difference divided by the standard deviation of the measure, so that magnitudes are comparable within and between panels; the number beside each point is the difference in the original units. Blue denotes p<0.05, grey no significant difference. Confidence intervals are given for every estimate in Table S2. Models adjusted for type of loss, gestational age, maternal age, years since loss and geographical area.

All women with a perinatal loss who answered the accommodation item are included (n=1769); the number contributing to each measure is given with the mean. Measures are scored 0 to 100 except respectful care and satisfaction with services (0 to 4), the Perinatal Grief Scale (11 to 162 in this sample) and the NSESSS (0 to 36).

These associations were not explained by ward type. Adding ward to the models left them substantially unchanged: grief 2.99 (95% CI 0.43 to 5.55), satisfaction with accommodation −28.71, respectful care −0.48, overall satisfaction −0.71 and the ward-environment section −19.87, with post-traumatic stress symptoms attenuated to 0.95 (95% CI −0.06 to 1.97, p=0.066). Interactions between accommodation and ward were not significant for grief (p=0.68), posttraumatic stress symptoms (p=0.48) or overall satisfaction (p=0.081), and weakly so for satisfaction with accommodation (p=0.041), where the advantage of separation was slightly smaller in gynaecology because the baseline there was already higher. Separation from newborns was therefore associated with better outcomes in both wards.

Women who shared accommodation with newborns were also less likely to have been able to stay with their partner (66.1% in the obstetric ward and 52.1% in gynaecology, against 83.5% and 75.5% where accommodation was separate), so shared accommodation marks a broader pattern of constrained care. The absolute rating given to the midwife was lower among women who shared (−11.22 after adjustment for ward), as was the rating given to the nurse, whereas the skill-mix differential between the two was unaffected by sharing (0.68, 95% CI −3.17 to 4.52, p=0.73). Shared accommodation is associated with care rated worse across the board, and it does not shift the composition of the team at the bedside.

### 3.3 How separation was achieved: gynaecology and midwifery presence

Among women for whom separation had been achieved, the gradient in midwifery presence did not follow separation alone. Women in the obstetric ward with separate accommodation reported the highest midwife ratings (78.7 for clinical care, 75.1 for emotional care) and the widest skill-mix differential in favour of the midwife (16.0). Women in a gynaecology ward with separate accommodation reported lower midwife ratings (66.9 and 63.6) and a much narrower differential (6.3), while their nurse ratings were essentially unchanged (60.7 versus 62.6). Table 2 shows the means by configuration, and Figure 2 the relation between midwifery presence and satisfaction with the accommodation.

**Table 2.** Midwifery presence, experience and psychological outcomes, by configuration of ward and accommodation.

| Measure | Obstetric,<br>separate | Obstetric,<br>shared | Gynaecology,<br>separate | Gynaecology,<br>shared |
| --- | --- | --- | --- | --- |
| *Midwifery presence, mean (95% CI)* |  |  |  |  |
| Midwife, clinical care | 78.7 (76.0, 81.3) | 58.0 (54.3, 61.7) | 66.9 (63.6, 70.2) | 51.4 (45.5, 57.4) |
| Midwife, emotional care | 75.1 (72.2, 78.0) | 50.8 (46.7, 54.9) | 63.6 (60.0, 67.2) | 44.0 (37.7, 50.3) |
| Skill-mix differential, clinical care | 16.0 (12.9, 19.2) | 12.1 (8.6, 15.5) | 6.3 (3.2, 9.5) | 2.6 (-2.4, 7.5) |
| Skill-mix differential, emotional care | 18.8 (15.2, 22.5) | 12.2 (8.4, 16.1) | 11.3 (7.9, 14.7) | 3.9 (-1.9, 9.8) |
| No midwifery care, % | 2.3 (n=477) | 5.2 (n=362) | 10.6 (n=461) | 12.0 (n=167) |
| *Other professionals, mean (95% CI)* |  |  |  |  |
| Nurse, clinical care | 62.6 (59.3, 65.8) | 44.1 (40.6, 47.7) | 60.7 (57.6, 63.7) | 48.6 (43.2, 54.1) |
| Nurse, emotional care | 56.4 (52.9, 59.9) | 35.6 (31.9, 39.3) | 51.3 (48.0, 54.6) | 41.2 (35.4, 47.0) |
| Obstetrician, clinical care | 64.0 (61.0, 67.0) | 47.3 (43.6, 51.0) | 61.1 (58.0, 64.2) | 53.0 (47.6, 58.5) |
| *Experience and outcomes, mean (95% CI)* |  |  |  |  |
| Satisfaction with accommodation (0-100) | 71.1 (68.6, 73.7) | 34.3 (31.1, 37.4) | 74.9 (72.5, 77.3) | 45.5 (40.8, 50.2) |
| Treated with respect (0-4) | 3.44 (3.35, 3.52) | 2.69 (2.53, 2.85) | 3.27 (3.16, 3.38) | 2.86 (2.56, 3.16) |
| Overall satisfaction with services (0-4) | 3.08 (2.96, 3.20) | 2.09 (1.91, 2.28) | 2.94 (2.81, 3.06) | 2.24 (1.89, 2.58) |
| Perinatal Grief Scale | 103.2 (101.2, 105.1) | 103.5 (101.1, 105.8) | 100.8 (98.7, 103.0) | 103.5 (99.6, 107.3) |
| NSESSS | 15.3 (14.5, 16.1) | 15.5 (14.6, 16.4) | 14.9 (14.1, 15.7) | 16.4 (14.9, 17.9) |
| *Check on the exposure variable* |  |  |  |  |
| CLASS ward-environment section | 78.5 (74.1, 82.8) | 53.9 (45.6, 62.1) | 78.0 (73.0, 83.0) | 45.9 (32.3, 59.5) |
| CLASS item, least disruptive ward | 84.0 (79.6, 88.5) | 58.3 (49.1, 67.5) | 84.5 (79.5, 89.4) | 47.9 (32.4, 63.4) |

**Figure 2.**
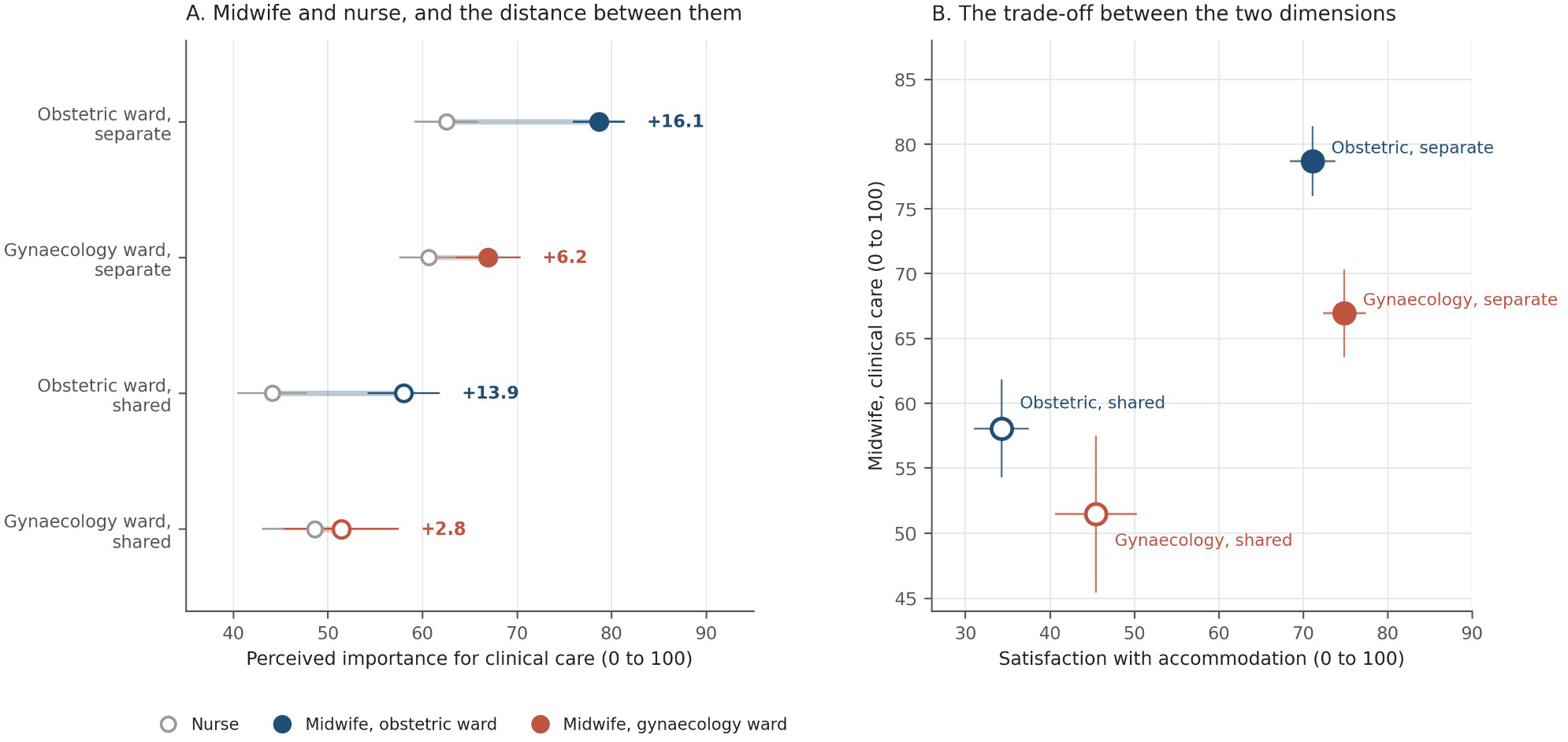
Panel A gives the mean perceived importance of the midwife and of the nurse for clinical care, with 95% confidence intervals; the segment joining them is the within-woman skill-mix differential, whose value is printed at the right. Panel B plots the midwife rating against satisfaction with accommodation, with intervals on both axes. Filled markers denote separate accommodation and open markers shared, blue the obstetric ward and red the gynaecology ward.

The skill-mix differential is the midwife rating minus the nurse rating within the same woman. No midwifery care denotes having rated the obstetrician but not the midwife. Adjusted differences between the two configurations in which separation had been achieved are shown in Figure 1 and in Table S2.

Both configurations had achieved separation from newborns, and adherence to the corresponding CLASS item was equally high in both (84.0 and 84.5), as was adherence on the whole ward-environment section (78.5 and 78.0). Women in gynaecology nonetheless reported midwifery clinical care 7.13 points lower (95% CI −11.18 to −3.09, p<0.001) and emotional care 6.89 points lower (95% CI −11.32 to −2.46, p=0.002) after adjustment, with a skill-mix differential 7.92 points narrower for clinical care (95% CI −12.47 to −3.36) and 6.54 narrower for emotional care (95% CI −11.50 to −1.57), and no difference in nurse ratings (0.74, 95% CI −3.89 to 5.37) or obstetrician ratings (−0.87). The proportion reporting no recognised midwifery care was 10.6% in gynaecology against 2.3% in the obstetric ward, an adjusted odds ratio of 3.05 (95% CI 1.49 to 6.23, p=0.002).

Satisfaction with accommodation ran the other way, and was higher in gynaecology by 7.33 points after adjustment (95% CI 3.84 to 10.82, p<0.001). Respectful care did not differ between the two separated configurations (−0.06, p=0.39), nor did overall satisfaction with services (−0.07, p=0.44), nor grief (0.55), nor post-traumatic stress symptoms (0.10). On one space-related checklist item, the assurance of parents’ privacy, gynaecology scored slightly higher (7.07, 95% CI 0.36 to 13.78, p=0.039), in line with the advantage in satisfaction with the accommodation.

### 3.4 Change over time and around the 2023 recommendations

Shared accommodation became steadily less common. The proportion sharing was 47.1% for losses before 2015, 44.1% for 2015 to 2019, 39.6% for 2020 to 2022 and 26.9% for losses from 2023 onwards, giving adjusted odds of 0.42 (95% CI 0.26 to 0.67, p<0.001) for the most recent period compared with the earliest. The step at 2023 was not simply the continuation of a smooth trend: in a model containing both, the indicator for losses from 2023 onwards retained an odds ratio of 0.72 (95% CI 0.54 to 0.94, p=0.016) alongside a per-year odds ratio of 0.961 (95% CI 0.927 to 0.996). Figure 3, panel A, shows the year-by-year series.

**Figure 3.**
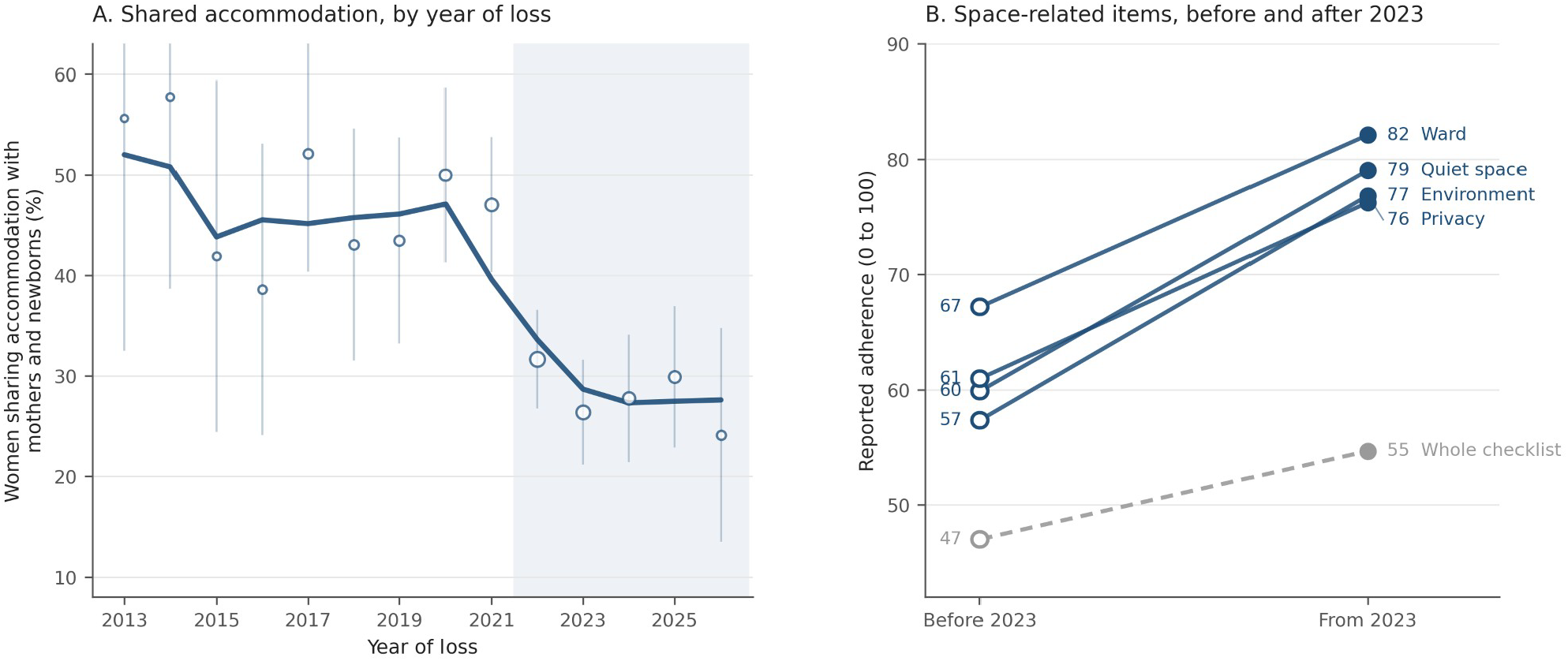
Panel A gives the proportion of women who shared their accommodation, by year of loss from 2013, with 95% Wilson intervals and marker size proportional to the number of women; the line is a three-year moving average weighted by that number, and the shaded area begins in 2022, when the recommendations circulated before their publication in February 2023. Panel B compares losses before 2023 with losses from 2023 onwards, restricted to stillbirth at 20 weeks or later and grouped by year of loss rather than by when the woman answered. The labelled items are the offer of the least disruptive ward and if possible a single room (Ward), a quiet and private environment for difficult conversations (Quiet space), the assurance of parents’ privacy (Privacy) and the ward-environment section (Environment); the whole 60-item checklist, as a percentage of its maximum, is the dashed grey line.

The space-related items of the checklist moved in the same direction and by a large margin. Within OPALE, adherence among women with losses from 2023 onwards was higher by 17.8 points on the item concerning the least disruptive ward, 23.3 on privacy, 19.3 on a quiet environment and 20.3 on the ward-environment section as a whole (all p<0.001). Among women with a stillbirth at 20 weeks or later, those whose loss occurred from 2023 onwards reported adherence higher by 14.9 points on the least disruptive ward (82.1 versus 67.2), 15.2 on privacy (76.2 versus 61.0), 19.1 on a quiet environment (79.0 versus 59.9) and 19.4 on the ward-environment section (76.8 versus 57.4), all p<0.001.

In the same years, the whole 60-item checklist mean score moved from 28.2 to 32.8 out of 60 (p<0.001), which on a 0 to 100 scale is a rise of about 8 points, against 19 points on the ward-environment section. The item on placing the baby in a cot in the room, which concerns the use of the room rather than its allocation, moved from 29.4 to 39.3 (p=0.043). Bereavement care improved overall, and it improved about two and a half times as much on the items that determine where the woman is accommodated (see Table S3 for details).

### 3.5 Sensitivity analysis at 20 weeks and later

Among the 963 women whose loss occurred at 20 weeks or later, the association between shared accommodation and outcomes persisted for grief (3.34, 95% CI 0.19 to 6.50, p=0.038), satisfaction with accommodation (−28.25), overall satisfaction with services (−0.68) and the ward-environment section (−18.64), and was attenuated for post-traumatic stress symptoms (0.99, p=0.12).

For the comparison between the two separated configurations, the advantage in satisfaction persisted (8.23, 95% CI 3.90 to 12.56, p<0.001) and the narrowing of the skill-mix differential persisted (−8.31, 95% CI −14.01 to −2.62, p=0.004), but the deficit in the absolute midwife rating attenuated and was no longer significant (−3.79, p=0.113), and the difference in no recognised midwifery care was no longer detectable (5 of 250 versus 1 of 317, Fisher’s exact p=0.092). At these gestational ages the woman gives birth, so a midwife is necessarily involved wherever she is admitted, and what changes is the relative weight of the midwife rather than her presence at all.

## 4. Discussion

A room away from mothers and newborns was associated with lower grief, fewer post-traumatic stress symptoms, markedly higher satisfaction with the accommodation, more respectful care and higher overall satisfaction with the services received. The associations survived adjustment for ward type and did not differ materially between the two wards. A recent review found no study that had measured the effect of bereavement space on grief or mental health,^5^ and these appear to be the first estimates of that effect.

Grief and post-traumatic stress are complex conditions and were assessed here months or years after the loss, so no single feature of a hospital admission should be expected to shift them by much.

Consistently, the differences were small, but significant: about three points on the Perinatal Grief Scale (ranging from 11 to 162), and about one point on the post-traumatic stress measure. Both survived multivariable adjustment. The measures of reported experience instead increased considerably, since they ask about the admission itself: 30 points out of 100 on satisfaction with the accommodation, and about half a point and three-quarters of a point on two four-point scales, roughly a fifth of the available range. The three sets of measures, grief, trauma and satisfaction, all point therefore the same way.

The general evidence on hospital accommodation is often invoked in support of single rooms and is weaker than assumed. Outside intensive care, reviews find no consistent clinical benefit, and a substantial minority of patients prefer shared accommodation because company protects them from isolation.^10^ The two literatures concern different exposures. In general wards the case for the single room rests on privacy, noise and dignity, and the countervailing risk is loneliness. After a perinatal death the exposure is different: the woman is placed among newborn babies while her own has died, and for most of them the company of other patients does not offset that.

Among women for whom separation had been achieved, those moved to a gynaecology ward were more satisfied with their accommodation but reported measurably less midwifery care, with three times the odds of reporting no midwifery care at all. The most plausible explanation is organisational: midwives are established in obstetric areas and deployed according to birth activity, gynaecology wards are staffed predominantly by nurses, and moving the woman changes the professional group at the bedside without any decision having been taken about her care needs. Nurse ratings did not fall when women were moved, obstetrician ratings did not fall either, and what fell was specifically the midwifery contribution. The workforce literature draws the same distinction.^13,14^Midwives describe bereavement care as demanding and resource-dependent,^24,25^ and early career midwives report feeling unprepared for it,^26^ so removing midwives from the pathway is unlikely to be neutral, and adding them without preparation would not be enough. A woman whose baby has died still needs the management of lactation, and Italian services deliver it unevenly: a quarter give no information about it after a loss, and follow-up is the component least often ensured after termination for fetal anomaly.^27,28^

Respectful care, overall satisfaction with services, grief and post-traumatic stress symptoms were indistinguishable between the two separated configurations, and satisfaction with the accommodation was higher in gynaecology. A service that audits its bereavement care by asking women whether they were satisfied and whether they were treated with respect, which is what most quality frameworks ask, would conclude that relocation had fully worked.

Shared accommodation became substantially less common across the period covered by this sample, falling from just under half of losses before 2015 to just over a quarter of losses from 2023 onwards, with a step at 2023 that persisted after allowing for a linear secular trend. On the space-related checklist items the improvement was between 15 and 19 points, against the equivalent of about 8 points for the checklist as a whole. Bereavement care improved on several fronts, and most where the recommendation can be met by changing an admission decision, without training, extra staff or a redesigned pathway. It is therefore likely to be acted on first, and the easiest way of acting on it is the one that costs midwifery presence. Anyway, we cannot causally attribute the change to the 2023 recommendations: the design is observational, the step and the recommendations coincide in time, and unmeasured change in maternity services is an equally plausible explanation.

Where the woman gives birth, at 20 weeks and beyond, a midwife is necessarily present, and relocation costs the relative prominence of midwifery care rather than its existence. Below that threshold, relocation to gynaecology can mean that the woman never encounters a midwife. These earlier losses are also the ones that international guidelines and the Italian 2023 recommendations largely do not cover, and in our data they were the losses most often spent alongside newborns: 44.6% after miscarriage, against 25.2% after stillbirth. These women are therefore the least covered by guidance, the most often accommodated alongside newborns, and the most likely to lose midwifery care when a service does move them.^18^

These data support separate accommodation, and they do so on measured outcomes rather than on principle alone: shared accommodation was the strongest correlate of poor experience and was associated with worse psychological outcomes. They also show that separation is not only a matter of space. The configuration that performed well on both dimensions was the separate room within the obstetric ward, which accounted for 546 of 1662 women, and separation had been achieved for 57.4% of women admitted to obstetric wards. Where the built environment allows, separation should be achieved within the maternity area rather than by moving the woman out of it. Where relocation is unavoidable, midwifery care should follow her rather than stay with the bed. Both depend on midwifery staffing and training being resourced, and both can be audited: services can count how many women were separated from newborns and how many were seen by a midwife.

### 4.1 Strengths and limitations

This analysis is large and national in scope, with harmonised geographical coding. Exposure was measured by a direct question rather than inferred, matched ratings were available for four professional groups within the same woman, and the pre-recommendation comparison comes from the same observatory, the same recruitment channels and the same checklist rather than from an outside source.

The design is cross-sectional and the sample is self-selected through the channels of a bereaved parents’ organisation, so it is not a probability sample of Italian women. Associations cannot be read causally.

The usual concern that memories fade with time carries less weight here: memories of traumatic events retain their coherence,^29^ the CLASS analysis found no difference in outcomes beyond twelve months from the loss once time was accounted for,^4^ and in the present data the association between accommodation and reported care was the same in women whose loss was within the past year and in those whose loss was earlier (interaction with years since loss, p=0.88 to p=0.97). What remains is that one questionnaire supplies both the exposure and the account of care, so a woman who recalls the admission more harshly may report both differently. The accommodation item asks whether the woman was in a room with mothers and their newborn babies, not how many beds the room had, so these results concern separation from newborns rather than single-room design; the exact percent of separated women who were in a single room was not recorded. The measure of midwifery presence is reported as a perception of importance rather than an objective record of staffing. Finally, this study is not an evaluation of any specific guideline, but it provides a baseline and two indicators implementation studies could track: the proportion of women sharing accommodation, which should fall, and the proportion reporting no midwifery care, which should not rise.

## Conclusion

In a national Italian sample, a third of women who experienced a perinatal loss spent their hospital admission alongside mothers and newborn babies. That exposure was associated with higher grief, more post-traumatic stress symptoms, substantially lower satisfaction with the accommodation and less respectful care. Shared accommodation also became markedly less common over the period studied, with a step coinciding with the 2023 national recommendations that was confined to the items about physical space. Where separation was achieved by moving the woman to a gynaecology ward, it came with a measurable reduction in midwifery presence and a threefold increase in the odds of receiving no midwifery care at all, most acutely in losses before 20 weeks, but that reduction did not appear in the woman’s own account of respect and overall satisfaction. Guidelines that require separate accommodation should also require that separation not remove the woman from midwifery care, and ask services to monitor both.

## Ethical statement

The OPALE study was approved by the Ethics Commission of the University of Florence (n. 175, prot. 0261728; n. 189, prot. 0333881). Participants received study information on the first page of the questionnaire and gave consent before proceeding. The survey was voluntary and anonymous, no personal identifiers were recorded, and the data were handled in accordance with Regulation (EU) 2016/679.

## CRediT authorship contribution statement

**Claudia Ravaldi:** Conceptualization, Methodology, Investigation, Writing – original draft, Writing – review and editing. **Alfredo Vannacci:** Methodology, Investigation, Formal analysis, Data curation, Software, Writing – original draft, Writing – review and editing.

## Declaration of generative AI in the scientific process

The authors used a large language model (Claude Opus 5, Anthropic) at two stages of this work. In the analysis, the design, the choice of measures and the specification of the models were conceived by the authors; the code implementing them was drafted and revised with the model, and its output was checked and validated by the authors against the source data. In the preparation of the manuscript, the model was used for drafting and editorial assistance, including polishing of academic English and formatting; all text was reviewed and, where needed, rewritten by the authors. The authors designed the study, verified every result reported here, and remain fully responsible for the content.

## Data availability

Aggregated data and analysis code are available from the corresponding author on reasonable request. Individual-level data cannot be shared because of the sensitivity of the information and the terms of consent.

## Supplementary material

Three supplementary tables follow the references. **Table S1** gives the characteristics of the 1662 women by configuration. **Table S2** gives every adjusted difference in numeric form, including the sensitivity analysis restricted to losses at 20 weeks or later. **Table S3** gives the space-related checklist items before and after the 2023 recommendations in women with a stillbirth at 20 weeks or later.

## Supplementary material

**Table S1.** Characteristics of the 1662 women, by configuration of ward and accommodation.

| Characteristic | Obstetric, separate<br>(n=546) | Obstetric, shared<br>(n=405) | Gynaecology, separate<br>(n=519) | Gynaecology, shared<br>(n=192) | p |
| --- | --- | --- | --- | --- | --- |
| Age at loss, years, mean (SD) | 34.9 (4.4) | 33.9 (4.8) | 34.5 (4.6) | 34.4 (4.8) | 0.014 |
| Gestational age, weeks, mean (SD) | 25.2 (9.8) | 21.6 (10.0) | 21.8 (10.3) | 18.6 (9.6) | <0.001 |
| Years since loss, mean (SD) | 1.2 (2.3) | 2.6 (3.9) | 2.0 (3.2) | 2.1 (2.7) | <0.001 |
| Type of loss, n (%) |  |  |  |  | <0.001 |
| Termination for fetal anomaly | 136 (24.9) | 78 (19.3) | 115 (22.2) | 39 (20.3) |  |
| Stillbirth | 243 (44.5) | 110 (27.2) | 201 (38.7) | 40 (20.8) |  |
| Neonatal death | 61 (11.2) | 76 (18.8) | 29 (5.6) | 23 (12.0) |  |
| Miscarriage | 106 (19.4) | 141 (34.8) | 174 (33.5) | 90 (46.9) |  |
| Educational level, n (%) |  |  |  |  | 0.995 |
| Secondary or below | 22 (4.0) | 13 (3.2) | 21 (4.0) | 8 (4.2) |  |
| Upper secondary | 161 (29.5) | 124 (30.6) | 153 (29.5) | 63 (32.8) |  |
| Bachelor or equivalent | 122 (22.3) | 86 (21.2) | 115 (22.2) | 37 (19.3) |  |
| University degree | 128 (23.4) | 104 (25.7) | 131 (25.2) | 45 (23.4) |  |
| Postgraduate | 113 (20.7) | 78 (19.3) | 99 (19.1) | 39 (20.3) |  |
| Macro-area of residence, n (%) |  |  |  |  | <0.001 |
| North | 338 (61.9) | 201 (49.6) | 328 (63.2) | 78 (40.6) |  |
| Centre | 130 (23.8) | 101 (24.9) | 104 (20.0) | 39 (20.3) |  |
| South | 53 (9.7) | 79 (19.5) | 65 (12.5) | 52 (27.1) |  |
| Islands | 25 (4.6) | 24 (5.9) | 22 (4.2) | 23 (12.0) |  |
| Previous perinatal loss, n (%) | 164 (30.0) | 142 (35.1) | 150 (28.9) | 70 (36.5) | 0.082 |

Separate denotes accommodation not shared with mothers and newborn babies. p values from linear regression across the four configurations for continuous variables and from Pearson chi-squared for categorical variables. Percentages are column percentages within each configuration.

**Table S2.**
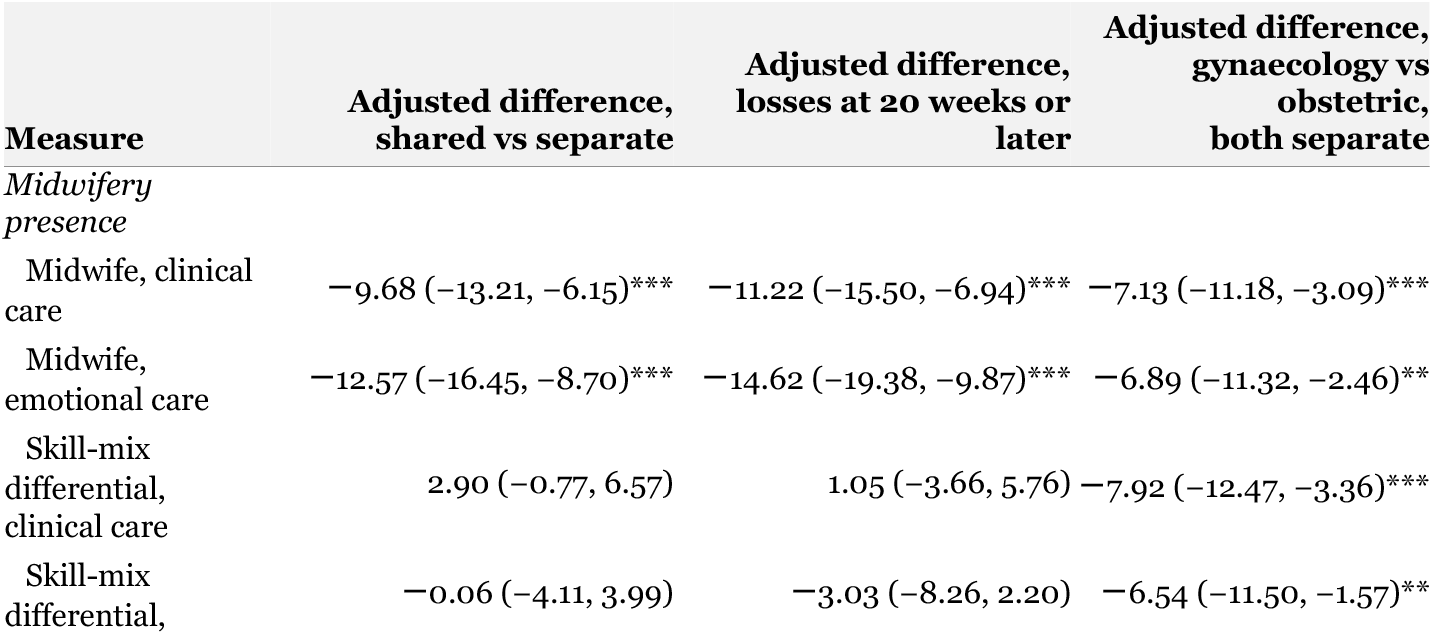
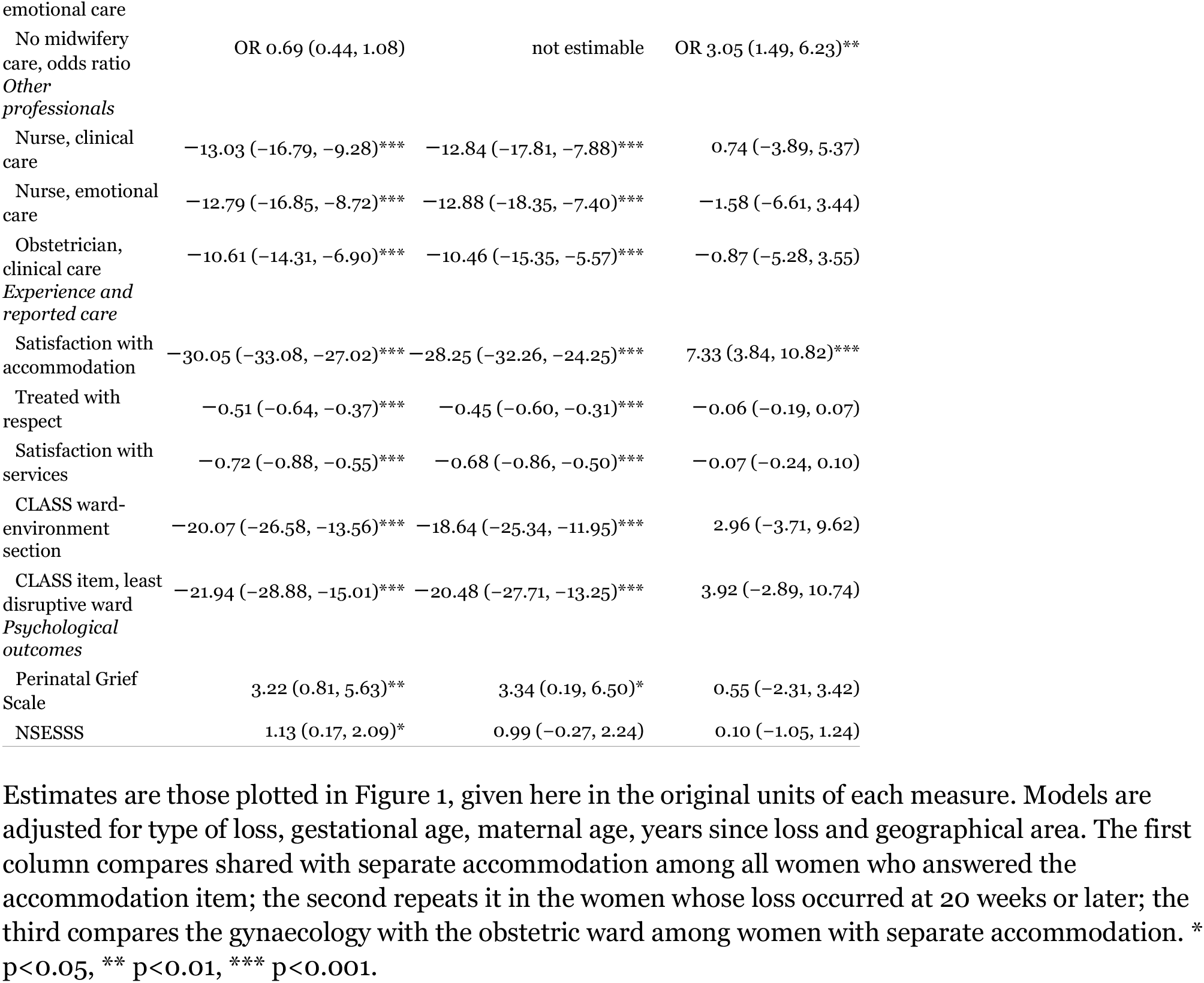
Adjusted differences for all measures, in numeric form.

**Table S3.** Space-related guideline items before and after the 2023 national recommendations, in women with a stillbirth at 20 weeks or later.

|  | Losses<br>before 2023 | Losses<br>from 2023 | p |
| --- | --- | --- | --- |
| Least disruptive ward, if possible a single room | 67.2 (62.8, 71.6) n=280 | 82.1 (77.8, 86.4) n=195 | <0.001 |
| Privacy of the parents | 61.0 (56.7, 65.2) n=284 | 76.2 (71.9, 80.6) n=205 | <0.001 |
| Quiet and private environment | 59.9 (55.7, 64.1) n=285 | 79.0 (74.9, 83.2) n=203 | <0.001 |
| Ward-environment section | 57.4 (53.4, 61.4) n=291 | 76.8 (72.8, 80.9) n=207 | <0.001 |
| Baby placed in a cot in the room | 29.4 (23.8, 35.0) n=211 | 39.3 (31.4, 47.1) n=117 | 0.043 |
| Whole 60-item checklist (out of 60) | 28.2 (26.3, 30.1) n=294 | 32.8 (30.9, 34.7) n=214 | <0.001 |

Women are grouped by the year of their loss and not by when they answered, so the earlier group includes both the women recruited first, whose responses supplied the CLASS validation analysis and whose losses all fall between 2008 and 2019, and the women recruited later who report a loss before 2023. Both groups are restricted to stillbirth at 20 weeks or later, the criterion of the earlier analysis, and both answered the same checklist within the same observatory. Values are means with 95% confidence intervals; p values from Welch t tests. The item on the cot in the room and the whole-checklist score are included as contrasts, to show that the change was concentrated in the items about where the woman is accommodated.

